# Comprehensive Risk Factors for Alzheimer’s Disease and Cognitive Function Before Middle Age in the U.S.

**DOI:** 10.1101/2024.10.31.24316509

**Authors:** Allison E. Aiello, Jennifer Momkus, Rebecca C. Stebbins, Yuan S. Zhang, Chantel L. Martin, Y. Claire Yang, Lauren Gaydosh, Taylor Hargrove, Adina Zeki Al Hazzouri, Kathleen Mullan Harris

## Abstract

**Importance:** Alzheimer’s disease (AD) is a major health concern in the U.S., but most research has focused on older populations. Few studies investigate AD risk factors and cognitive function in young to early midlife adults.

**Objective:** To examine whether key AD risk factors are associated with cognition before midlife.

**Design, Setting, Participants:** Data from the National Longitudinal Study of Adolescent to Adult Health (Add Health) were analyzed. Participants were enrolled in 1994-95 (grades 7-12) and followed through 2018. We analyzed survey and biomarker data from Waves IV (median age 28) and V (median age 38).

**Exposures:** Cardiovascular Risk Factors, Aging, and Incidence of Dementia (CAIDE) score, APOE ε4 status, Amyloid Tau and Neurodegeneration markers (ATN), including total Tau and Neurofilament light (NfL), high sensitivity C-reactive protein (hsCRP), Interleukin (IL)-1β, IL-6, IL-8, IL-10, and Tumor necrosis factor alpha (TNF-α).

**Main Outcomes:** Immediate word recall, delayed word recall, and backward digit span.

**Results:** We analyzed data separately in Wave IV (ranging from N=4,507 to N=11,449) and Wave V (ranging from N=529 to N=1,121). Approximately half were female. The CAIDE score was associated with all cognition measures in Wave IV. For example, among adults aged 24-34, each 1-point increase in CAIDE was associated with a 0.03 SD lower backward digit span score (95% CI: −0.04, −0.02). No significant associations were found between APOE ε4 and cognition. Total Tau was associated with immediate word recall in Wave V (β=-0.14, 95% CI: −0.24, −0.04). Wave IV hsCRP and IL-10 and Wave V IL-6, IL-1β and IL-8 were also associated with lower cognitive scores.

**Conclusions:** Key risk factors for AD, including cardiovascular, ATN, and immune markers, are linked to cognitive function as early as ages 24-44, highlighting the need for early prevention in the US.

## Background

Alzheimer’s disease (AD) poses a global health challenge.^1^ While most research in the United States (US) has focused on older populations, where the risk of dementia is highest, there is a lack of studies investigating risk factors for AD and cognitive function in young to early midlife adults before the onset of disease. Identifying pathways to AD and cognitive impairment before older age is critical for slowing the expected growth of AD in coming decades.

The landscape of risk factors that are known to shape AD risk in older age encompass social, behavioral, clinical, and biological dimensions.^1^ On the social and behavioral side, factors such as lower educational attainment, physical inactivity, and smoking are notable risk factors. Clinically, health conditions such as diabetes, obesity, hypertension, and high cholesterol are identified as consistent contributors to AD risk. In recent years, researchers have sought to combine the social, behavioral, and clinical risk factors into comprehensive risk scores for AD.^2^ The Cardiovascular Risk Factors, Aging, and Incidence of Dementia (CAIDE) score is among the most extensively researched risk algorithms and integrates social, behavioral, and biological risks including education, sex, age, cholesterol levels, blood pressure, body mass index, and physical activity.^3^

On the biological side, genetic, neurological, immune, and inflammatory molecules have been implicated as biomarkers of AD risk.^4^ The genetic profile of apolipoprotein E (APOE) is a known risk factor for AD in older populations.^5^ Moreover, neuropathological markers, such as amyloid beta and neurofibrillary tangles, have been investigated as predictors of AD in later life. The amyloid (A), tau (T), and neurodegeneration (N) (known as ATN) biomarkers are increasingly promising for predicting AD risk in older populations.^6^ Two commonly measured biomarkers are total tau and neurofilament light chain (NfL). Both measures are predictive of later life AD risk in older populations.^7,8^ Furthermore, central and peripheral immune and inflammatory mechanisms are increasingly recognized as pivotal in the pathogenesis of AD. A recent comprehensive review identified relatively consistent positive associations between several inflammation-related cytokines and AD risk, with the two most-studied markers being interleukin-6 (IL-6) and tumor necrosis factor-alpha (TNF-α).^4^ While also widely studied, C-reactive protein was less consistently associated with AD risk in this review, with approximately 25% of the studies demonstrating positive associations, 25% showing negative associations, and roughly 50% showing no significant association. Less well-studied candidate immune and inflammatory markers have been examined in relation to AD, such as IL-1β and IL-8, but the small body of existing studies showed inconsistent findings.^4^

While most studies analyzing CAIDE risk scores, ATN, and immune biomarkers have focused on older populations, their relationship with cognitive function in pre-midlife adults has rarely been investigated. This study aims to address several significant gaps in our understanding of AD risk in the US population by examining widely studied risk factors for AD and cognitive function before midlife. This is crucial because many have hypothesized a long subclinical phase of AD but have rarely studied AD markers in early adulthood. Specifically, we assessed the associations between CAIDE score, APOE ε4 status, two ATN biomarkers (total Tau and NfL), and several inflammatory molecules and interleukins (hsCRP, IL-1β, IL-6, IL-8, IL-10, TNF-α) with three standard measures of cognitive function in a US representative study of 24- to 44-year-old participants in the National Longitudinal Study of Adolescent to Adult Health (Add Health).^9^ Our findings aim to shed light on the emergence of AD risk in early adulthood in the U.S. population over the forthcoming two to four decades.

## Methods

### Study Population

Data from Waves IV-V of the National Longitudinal Study of Adolescent to Adult Health (Add Health) were used for this study. Add Health is a nationally representative cohort tracking adolescent participants since 1994-1995 (Wave I) through four follow-up waves.^9^

Wave IV (n=15,701), conducted in 2008, when participants were 24-34 years old, included in-home interviews, cognitive tests, physical exams, saliva collection, and dried blood spot collection from all eligible respondents. Wave V, conducted between 2016 and 2018 when participants were aged 34-44, was a mixed-mode survey combining in-person and web/mail surveys with a target of 12,300 survey participants, out of which 1,702 received in-home interviews. Cognitive tasks were administered in-person by field interviewers during the in-home interviews. All Wave V participants were invited to complete a separate “biovisit”, including a physical exam and venous blood collection, resulting in visits arranged for 5,381 participants. Additional information on study design, response rates, sample collection, and testing procedures are available at Harris et al. 2019 and on the Add Health website.^9,10^ Participants who reported being pregnant were removed from analyses except those involving APOE status only.

### Measures

#### CAIDE Score

The CAIDE score is a weighted composite score based on established risk factors for dementia. There are two versions; both include age, education, biological sex, systolic blood pressure (SBP), body mass index (BMI), total cholesterol, and physical activity. Version 2 (v2) adds APOE ε4 status. The weights are the coefficients derived from the original study, creating the CAIDE score.^3^ We aimed to align as closely as possible with the variables used in the original study while adapting to our specific sample and available data. A summary of the components and scoring are shown in Table 1.

**Table 1.** CAIDE Score Components.

| CAIDE Risk Factors | Version 1 Weighted Components | Version 2 Weighted Components |
| --- | --- | --- |
| Age | Lowest tertile = 0<br>Middle tertile = 3<br>Highest tertile = 4 | Lowest tertile = 0<br>Middle tertile = 3<br>Highest tertile = 5 |
| Educational Attainment | College degree or higher = 0<br>Some college/technical training = 2<br>HS diploma/GED or less = 3 | College degree or higher = 0<br>Some college/technical training = 3<br>HS diploma/GED or less = 4 |
| Biological Sex | Female = 0<br>Male = 1 | Female = 0<br>Male = 1 |
| Systolic Blood Pressure | $\leq 140$ mm Hg = 0<br>>140 mm Hg = 2 | $\leq 140$ mm Hg = 0<br>>140 mm Hg = 2 |
| BMI | $\leq 30$ kg/m<sup>2</sup> = 0<br>>30 kg/m<sup>2</sup> = 2 | $\leq 30$ kg/m<sup>2</sup> = 0<br>>30 kg/m<sup>2</sup> = 2 |
| Total Cholesterol | Bottom 9 deciles = 0<br>Top decile = 2 | Bottom 9 deciles = 0<br>Top decile = 1 |
| Physical Activity | ≥ activity 2x per week = 0<br>< activity 2x per week = 1 | ≥ activity 2x per week = 0<br>< activity 2x per week = 1 |
| APOE e4 status | N/A | Non-$\epsilon 4$ carrier = 0<br>$\leq 1$ $\epsilon 4$ allele = 2 |

#### APOE ε4

A total of N=11,550 participants had available genomic data. The Michigan Imputation Server was used for imputation on unphased data. Data were imputed using 1000 Genomes Project phase 3 version 5.^11^ Using the imputed rs7412 (R^2^ score = 0.896) and rs429358 (R^2^ score = 0.919) variants, we categorized participants as ε2/ε2, ε2/ε3, ε2/ε4, ε3/ε3, ε3/ε4, and ε4/ε4. APOE status was defined by having at least one ε4 allele (i.e. those with APOE ε2/ε4, ε3/ε4, or ε4/ε4 phenotypes vs. ε2/ε2, ε3/ε3, or ε2/ε3).

Amyloid, Tau, and Neurodegeneration (ATN) Biomarkers. Among those who completed a separate biovisit in Wave V, N=4,940 venous blood samples were collected via phlebotomy. Using a digital enzyme-linked immunosorbent assay (ELISA), serum samples were assayed for NfL, and plasma samples for total Tau.

#### Immune Biomarkers

Among Wave IV respondents, 75.9% consented to contribute a dried blood spot (DBS) sample and to sample archival (N=11,917). DBS samples were assayed using a high-sensitivity ELISA for CRP (hsCRP) in DBS samples^12^ and subsequently archived. A subset of archived DBS samples with sufficient quantity (N=5,019) were later tested for additional inflammatory markers, including interleukins (IL-6, IL-8, and IL-10) and TNF-α, using a highly sensitive electrochemiluminescent immunoassay (ECLIA) modified for DBS samples.^13^ In Wave V, serum samples from the biovisit (N=4,940) were assayed for hsCRP immediately upon receipt at the testing laboratory using a hsCRP-specific particle-enhanced immunonephelometric assay.

Archived samples were later tested for inflammatory cytokines (IL-6, IL-10, IL-8, IL-1β, and TNF-α) using an ECLIA.^14^ Units for all blood biomarkers are in pg/mL except for hsCRP (mg/L). Values were log-transformed then standardized.

#### Cognitive Function

Home interviewers conducted cognitive assessments among all participants in Wave IV (N=14,732) and those who received an in-home interview in Wave V (N=1,717). In each wave, three cognitive assessments included immediate word recall, delayed word recall, and backward digit span. Participants were read a 15-word list for word recall and asked to recall as many of the 15 words as possible.. Cognitive scores were standardized into a z-score for analysis, with a higher score indicating higher memory cognition.

#### Covariates

We identified covariates for inclusion based on Directed Acyclic Graphs (DAG) models.^15^ Early life SES was operationalized using the social origins score, based on parental education, occupation, household income, and receipt of public assistance.^16^ Additional covariates include age at exam, educational attainment (at the same wave in which the biomarker was measured), sex assigned at birth, and an indicator for recent inflammatory conditions. A race/ethnicity variable was constructed from the Wave V survey and supplemented with Wave I data if missing. Participants self-selected one or more boxes from a list including: “American Indian or Alaska Native”, “Asian”, “Black, African American”, “Hispanic”, “Pacific Islander”, “White”, and “Some other race or origin”. Please see Supplement for more information on variables and conceptualization of race and ethnicity.

#### Analysis

The independent variable, dependent variable, and sample size of each analytic sample are shown in Table 2. Flow charts illustrating the selection of each sample are provided in the supplementary material. Survey-weighted descriptive statistics were compiled to characterize demographics and relevant study variables in each sample. To examine the cross-sectional associations between cardiovascular (e.g., CAIDE score), ATN, immune, and genetic risk with cognitive function, survey-weighted linear regressions were used to estimate β parameters for unadjusted and adjusted models with each independent variable predicting each participants cognitive score in the same wave. For the CAIDE score models, covariates included race/ethnicity categories, social origins score, and an indicator for inflammatory conditions. For the ATN and immune BB biomarker models, covariates included sex assigned at birth, race/ethnicity categories, educational attainment, age, and an indicator for recent inflammatory conditions. For the APOE status, covariates included age and sex assigned at birth.

**Table 2.**
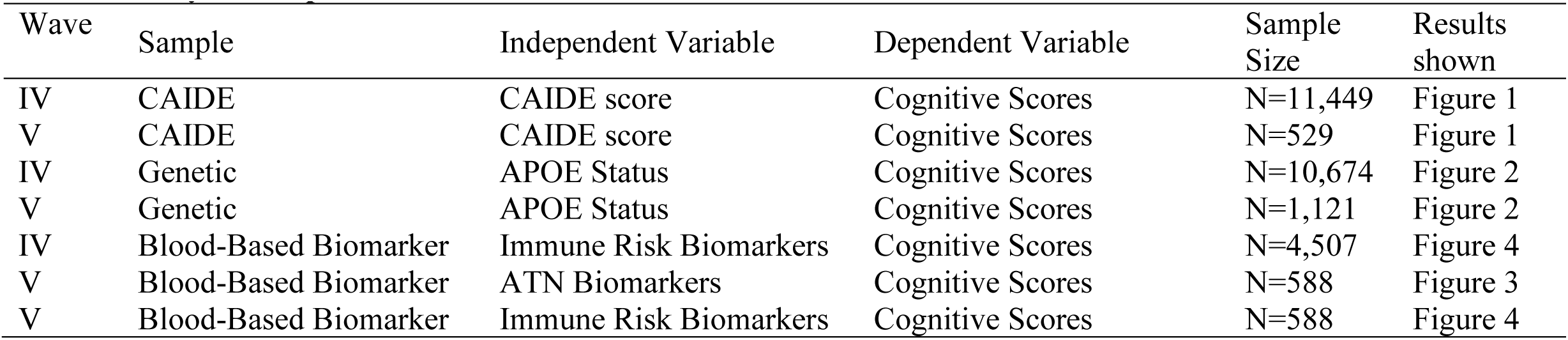
Analytic Samples.

## Results

The weighted characteristics of each analytic are shown in Table 3, and a comparison with the overall Add Health sample is available in Table S2. In Wave IV, the analytic sample sizes ranged from N=4,507 for the immune risk factors to N=11,449 for the CAIDE score, with a median age of 28 years. About half of the participants in Wave IV were female (48-52% across analytic samples), and about a third had a college degree or higher (26-30%). Approximately 70% were White (71-72%), 13-15% were Black, and 9-10% were Hispanic. In Wave V, analytical samples ranged from N=529 to N=1121, with a median age of 38. Approximately 45% were female (44-47%), and over a third had a college degree (33%-36%). Approximately 69% were White (68-69%), 20% were Black (17-20%), and 7-8% were Hispanic in Wave V.

**Table 3.**
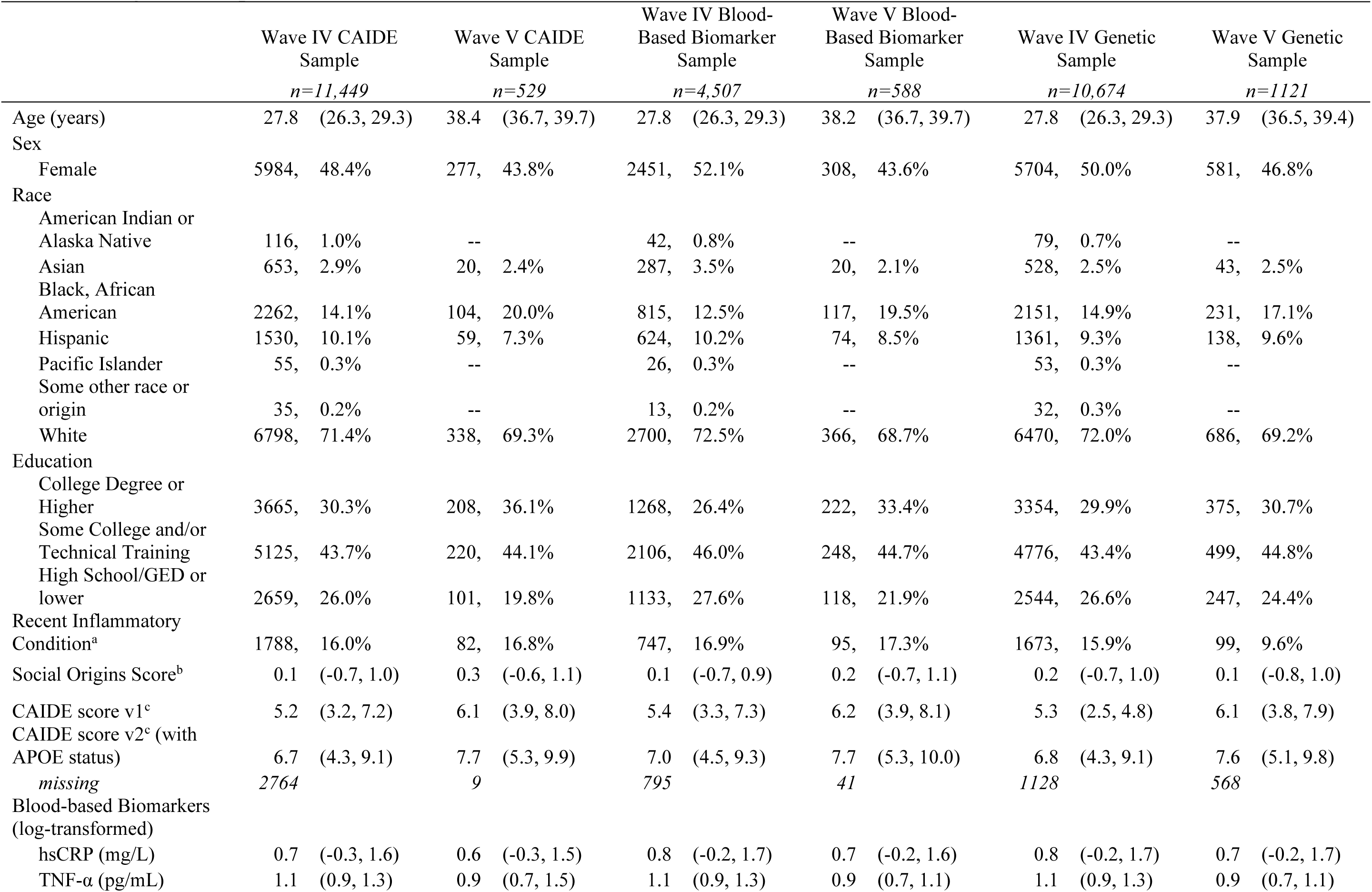
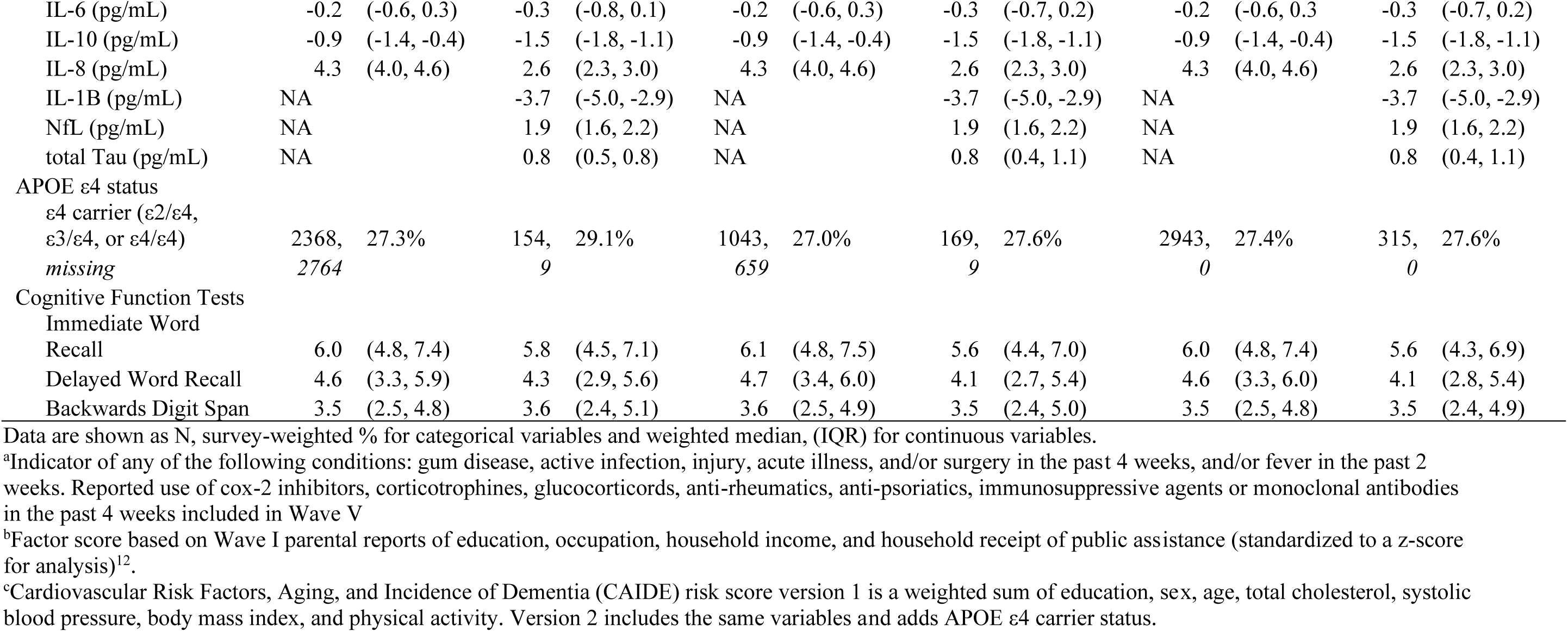
Weighted Sample Characteristics, Add Health Wave IV-V.

### CAIDE scores

In Wave IV, higher CAIDE scores (both versions) were statistically significantly associated with lower cognitive scores in all domains (Figure 1 and Table S3). At Wave V, higher CAIDE scores trended in the same negative direction as lower cognitive scores across measures, but the associations were not statistically significant (Figure 1 and Table S3). In supplementary analyses, we explored associations between CAIDE scores and cognitive measures across participants who had data on all variables and covariates in both Wave IV and Wave V (Supplementary Figure S3 and Table S4, N=412). Similar patterns were observed for both Waves IV and V, but some results became non-significant with reductions in sample size.

**Figure 1.**
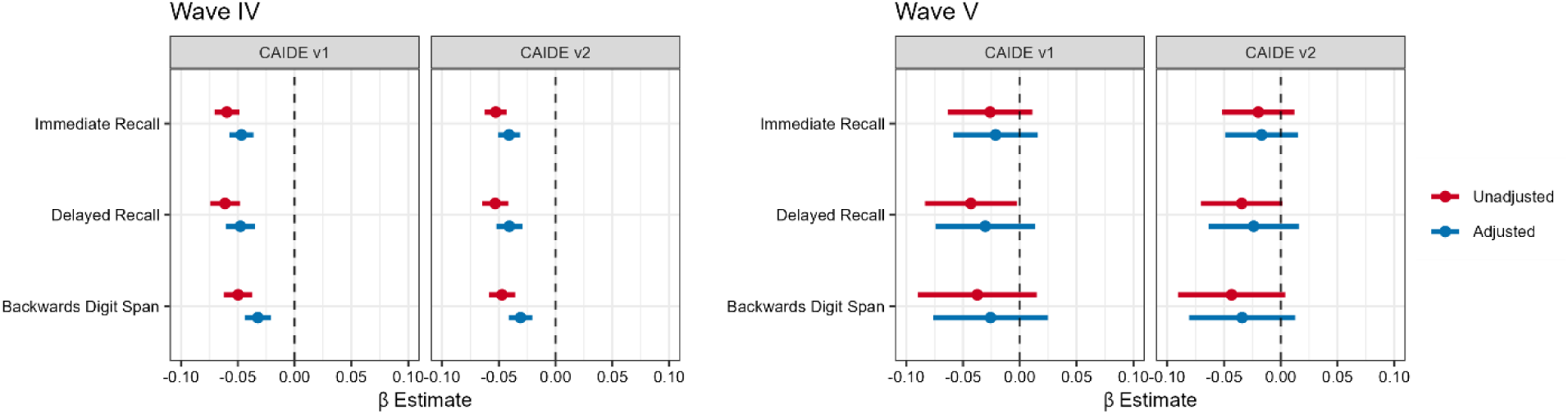
Association between CAIDE Score and Cognitive Test Scores, Add Health Wave IV and Wave V. Each panel shows the β estimates and 95% CIs for survey-weighted linear regressions where CAIDE scores are the independent variable and cognitive function tests scores are the dependent variable. Models are adjusted for race/ethnicity, social origin score, and an indicator for recent inflammatory conditions. Wave IV: N=11,449 for the CAIDE v1 score, N=8,685 for the CAIDE v2 score (with APOE e4 carrier status). Wave V: N=529 for CAIDE score v1, N=520 for the CAIDE score v2.

### Apolipoprotein (APOE) ε4

There were no associations between having at least one ε4 allele and cognitive function in Wave IV nor Wave V (Figure 2 and Table S3). We also examined the same associations among participants with data on all variables and covariates in both Wave IV and V (Supplementary Figure S3 and Table S4, N=1063). In this subsample, there were similarly no associations between APOE status and cognitive function.

**Figure 2.**
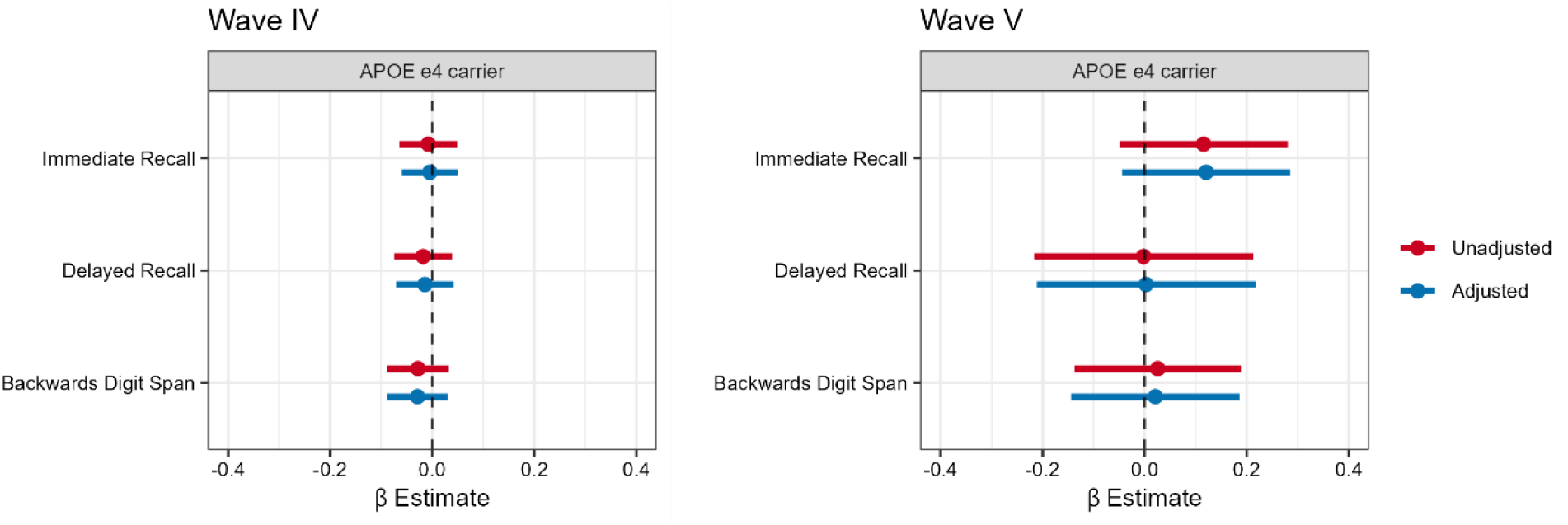
Association Between APOE Status and Cognitive Function, Add Health Wave IV and Wave V. Each panel shows the β estimates and 95% CIs for survey-weighted linear regressions where APOE e4 carrier status (carrier vs. non-carrier) is the independent variable and cognitive function tests scores are the dependent variable. Models adjusted sex assigned at birth and age. Wave IV N=10,674 Wave V N=1,121

### ATN markers

There was a negative association between total Tau and immediate word recall at Wave V shown in Figure 3 and Supplementary Table S3 (β=-0.14, 95% CI: −0.24, −0.04), but not with delayed recall nor the backward digit span. Nfl was associated with lower backward digit span scores, though it was not statistically significant (β=-0.08, 95% CI: −0.17, 0.01). There were no statistically significant associations between Nfl and the other cognitive measures.

**Figure 3.**
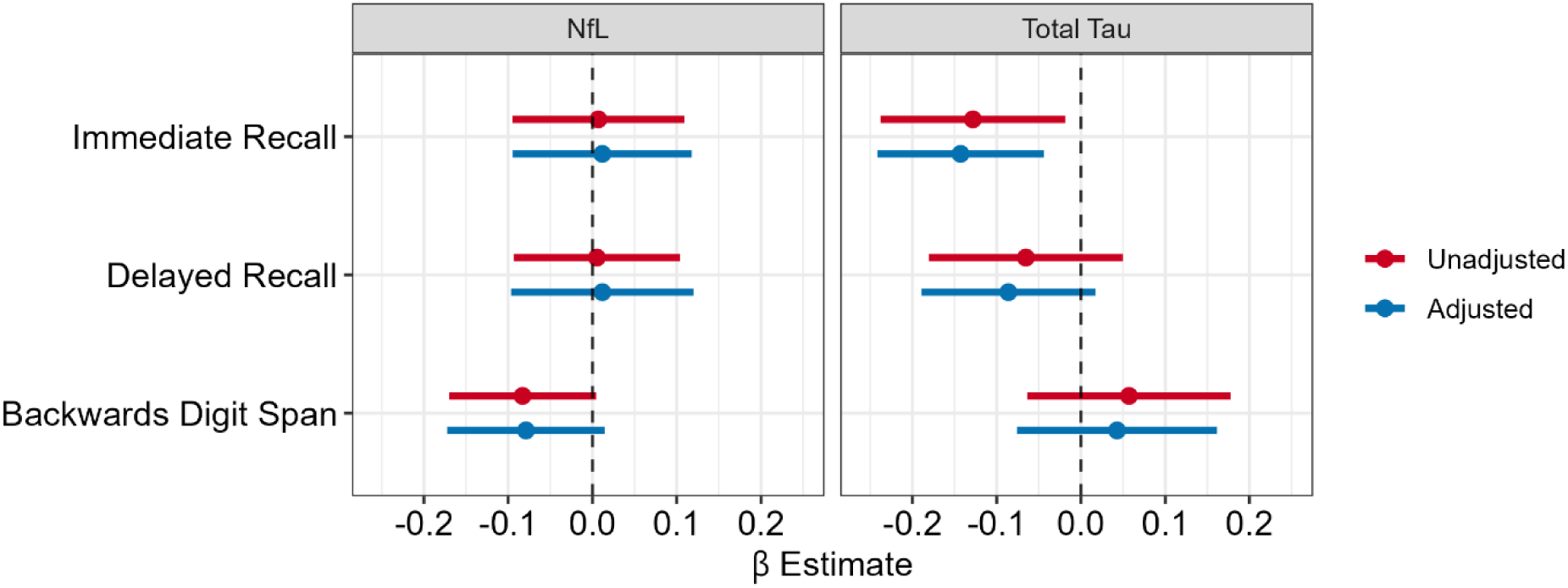
Association Between Amyloid, Tau, and Neurodegeneration (ATN) Biomarkers and Cognitive Function, Add Health Wave V, N=588. Each panel shows the β estimate and 95% CI for survey-weighted linear regressions where Wave V blood-based ATN biomarker concentrations are the independent variable and Wave V cognitive function tests scores are the dependent variable. Models adjusted for race/ethnicity, education, sex assigned at birth, age, and an indicator for inflammatory conditions. NfL N=588, total Tau N=584

### Immune risk factors

Cross-sectional associations between each inflammatory biomarker and cognitive scores are shown in Figure 4. β estimates are also shown in Supplementary Table S3. In Wave IV, IL-6 and hsCRP were associated with lower backwards digit span scores, but the estimate for IL-6 was not statistically significant after adjusting for covariates (IL-6: β=-0.04 SDs, 95% CI:-0.08, 0.00, hsCRP: β= −0.04, 95% CI: −0.08, −0.002). IL-10 was associated with delayed recall scores (β= −0.05, 95% CI: −0.09, −0.01). None of the other biomarkers (TNF-α, IL-8) were associated with lower cognitive scores in Wave IV (Table S3). In Wave V, IL-6 was associated with lower backward digit span (β=-0.10, 95% CI: −0.19, −0.01), and IL-8 was associated with lower cognitive scores in all domains (immediate recall: β=-0.18, 95% CI:-0.28, −0.09; delayed recall: β=-0.17, 95% CI:-0.29, −0.05; backward digit span: β=-0.19, 95% CI:-0.29, −0.09), even after adjustment for covariates. IL-1β was associated with lower immediate (β=-0.15, 95% CI:-0.28, −0.03) and delayed recall scores (β=-0.12, 95% CI:-0.21, −0.02). See Table S3 in the supplementary material for β estimates and 95% CIs of all the associations shown in the figures.

**Figure 4.**
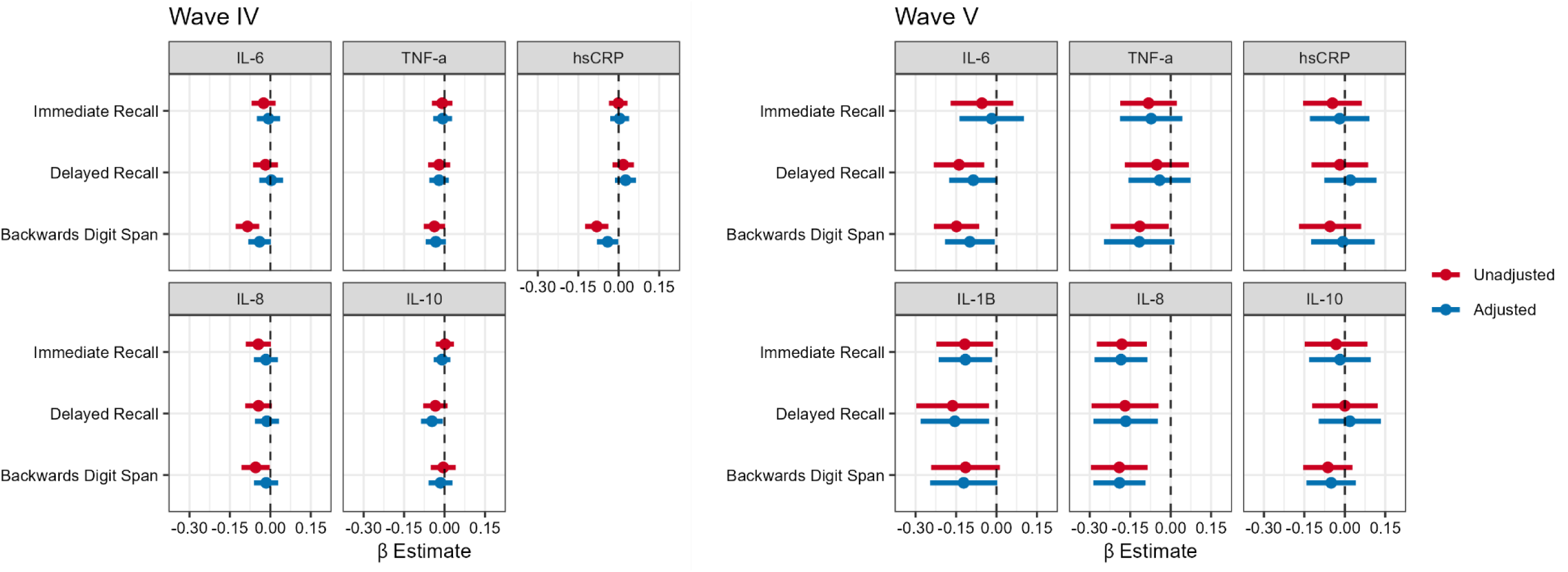
Association between Immune Risk Biomarkers and Cognitive Test Scores, Add Health Wave IV and Wave V. Each panel shows the β estimates and 95% CIs for cross-sectional survey-weighted linear regressions where inflammatory biomarker concentrations are the independent variable and cognitive function tests scores are the dependent variable. Wave IV cross-sectional associations are on the left and Wave V cross-sectional associations are on the right. Models adjusted for race/ethnicity, educational attainment, sex assigned at birth, age, and an indicator for inflammatory conditions Wave IV (dried blood spots): N=4485 for hsCRP, N=4507 for all other biomarkers Wave V (serum): N=567 for hsCRP, N=588 for all other biomarkers.

## Discussion

Several key risk factors for AD were associated with standard measures of cognition in 24- to 44-year-olds in the U.S., suggesting that these factors may be related to cognitive function decades before the onset of AD. The CAIDE score was consistently linked to all three cognitive function measures (immediate recall, delayed recall, and backward digit span). Notably, a key genetic risk factor, APOE ε4, was not associated with recall nor backward digit span, suggesting that the effects of APOE ε4 may not become apparent until middle to older age. We also observed significant associations with ATN biomarkers. Specifically, total Tau was associated with lower immediate word recall but not backward digit span scores. Although higher levels of NfL showed a trend toward lower backward digit span scores, the association was not statistically significant. A limited number of immune markers were significantly associated with cognitive function at ages 24-34 years; however, some of these associations appeared to become more robust in the following decade of life. For instance, IL-6 was not associated with backward digit span in Wave IV but showed a significant association with lower backward digit span in Wave V. Similarly, IL-8 was not associated with cognitive scores in Wave IV but was associated with all three measures of cognition in Wave V. These findings suggest that cardiovascular, ATN, and immune markers may be associated with critical measures of cognitive function at much younger ages than previously recognized, with some associations becoming more prominent between the early midlife ages of 33 and 44.

We observed strong associations between the CAIDE score and all three measures of cognition. For example, among adults aged 24-34, each 1-point increase in the CAIDE score was associated with a 0.03 SD decrease in the average backward digit span score, adjusted for covariates (95% CI: −0.04, −0.02). Among adults aged 34-44, each 1-point increase in the CAIDE score was similarly associated with a 0.03 SD decrease in the average backward digit span score, adjusted for covariates (95% CI: −0.08, 0.02). The magnitudes of these associations were consistent across waves for each cognitive test. Still, they did not reach statistical significance in Wave V. It is possible that sample size in Wave V was too small to detect statistically significant associations. Prior research supports robust associations between the CAIDE score and cognitive function in older cohorts, although these relationships varied depending on the cognitive domain and study population. For instance, studies in a US cohort (mean age 60.1 years) and a Finnish cohort (mean age 52.4 years) found consistent associations between higher CAIDE scores at baseline and lower subsequent scores in all domains tested (including memory tasks).^17,18^ In a New Zealand cohort, researchers identified an inverse association between the CAIDE score and midlife IQ and subjective midlife cognitive problems at age 45.^2^ However, a study in Northern Manhattan (mean age 64 years) found cross-sectional inverse associations between the CAIDE score and tests of executive function and processing speed, but no association with memory tests or decline in test scores over time.^19^ Finally, a study in the UK (mean age 56 years) showed associations between the CAIDE score and declines in reasoning, vocabulary, and global cognitive scores but not in memory scores.^20^ Importantly, all these studies focused on midlife or older ages (i.e., the average ages in each sample were above 45 years at baseline). To the best of our knowledge, no studies have examined the relationship between the CAIDE score and cognition in young adulthood and early midlife in the U.S. Our study contributes to this body of knowledge by suggesting that these significant associations are observable well before age 50.

In contrast to prior research on APOE ε4 and cognitive function in older populations, we did not observe significant associations between the APOE ε4 allele (heterozygous nor homozygous) and our measures of cognitive function. Our findings are consistent with previous research showing that the impact of APOE ε4 is not evident in younger populations.^21,22^ Taken together, these results support the hypothesis that tau-related pathology in conjunction with APOE ε4 may either accumulate gradually after age 50 or activate more rapidly in older age.^21,22^ Future research tracking these associations in longitudinal studies, such as Add Health, may help uncover the critical temporal changes in cognition related to APOE ε4.

Of note, two key ATN biomarkers appear to be associated with different aspects of memory at ages 34-44. NfL was associated, though not statistically significantly, with lower backward digit span scores, a measure of working memory. In contrast, total Tau was significantly associated with poorer immediate word recall, a measure of verbal episodic memory. Remarkably, these patterns are consistent with research on these markers in much older populations at risk of AD.

For example, NfL, a marker of neuronal injury, is particularly associated with working memory and processing speed.^23^ A study involving older adults from the National Health and Nutrition Examination Survey (NHANES) reported an inverse relationship between serum NfL levels and digit symbol substitution test (DSST) scores in older participants.^23^ Our finding of an association between Tau levels and episodic memory is noteworthy because Tau has been shown to be cross-sectionally associated with worse episodic memory, particularly among older individuals, including those without significant cognitive impairment.^24^ Together, these findings provide preliminary evidence that ATN biomarkers may influence episodic and working memory a decade before middle age. To our knowledge, these are the first findings to suggest that ATN biomarkers are associated with cognition in a relatively large sample of 34-44-year-olds. Future research is warranted to examine these and other promising ATN biomarkers in younger individuals.

We observed that several immune markers were associated with cognitive measures at ages 24-34, with IL-6 and hsCRP associated with backward digit span. In the next decade of life, IL-6 continued to be associated with backward digit span, while IL-1β and IL-8 appeared to be associated with word recall and backward digit span in Wave V but not Wave IV. In contrast, we found no associations with hsCRP in Wave V. Chronic systemic inflammation, often measured by elevated levels of inflammatory biomarkers like CRP and IL-6, has been linked to an increased risk of developing AD.^25^ A recent review by Li et al. found that IL-6 was the most consistently associated biomarker with AD, and IL-1β was also consistently positively associated with AD.^4^ However, these studies were primarily conducted in populations over the age of 50 years. Our findings suggest the potential emergence of IL-6 and IL-1β as important immune molecules related to cognition early in the life course, consistent with this review of later-life relationships. Our evidence for these immune and inflammatory relationships as early as the third and fourth decades of life, as observed in this study, is noteworthy and suggests that associations between these markers and cognition are apparent 10-20 years earlier than what prevailing theories suggest.^26^

This study has several limitations. Significant associations with CRP and IL-10 that we observed in Wave IV were not observed in Wave V, and may be related to the smaller sample size in Wave V. The limited sample size of the longitudinal data, precluded us from examining trends over time and ATN biomarkers were only available in Wave V. Wave IV samples were assayed for cytokines using blood spots, while Wave V samples were assayed using serum or plasma. As previously reported, IL-8 in particular, showed lower agreement between blood spot and serum assays.^13^ This discrepancy might explain why we observed an association between IL-8 and cognition in Wave V but not in Wave IV, which used blood spot assays. Nonetheless, this study is groundbreaking because it is the first to examine key AD risk factors in a large diverse population-based sample of 24- to 44-year-olds in the US.

In conclusion, our study highlights that crucial risk factors for AD, including cardiovascular, ATN, and immune markers, are associated with cognitive function much earlier in life than previously recognized, with significant associations observed as early as ages 24-44. The CAIDE score, in particular, consistently predicted cognitive scores across multiple domains in young adults, while APOE ε4’s impact may not emerge until later in life. ATN and immune and inflammatory biomarkers are already beginning to show associations with cognition pre-midlife.

Our findings suggest that key risk factors for AD in older age, begin to shape cognitive function decades before clinically detectable cognitive impairment and AD, underscoring the importance of early prevention across the life course.

## Data Availability

All data produced in the present study will be available to Add Health Users in 2026.

