## Supplement for "Comprehensive Risk Factors for Alzheimer’s Disease and Cognitive Function Before Middle Age in the U.S."

**Supplementary Material**

### Additional information on Variables

Components of the CAIDE score**:** Age at the time of the survey were categorized into tertiles (in months plus years) within the entire wave. The highest tertile, middle, and lowest tertile received a weighted score of 4, 3, and 0, respectively. Educational attainment is based on survey responses at each wave and categorized as high (college degree or higher, score=0), medium (some college and/or technical training, score=2), and low (high school diploma/GED or lower, score=3). Males received a score of 1, while females received a 0. SBP measurements were taken by trained personnel during the in-person exam. Following current guidelines, we averaged the second and third SBP measurements. Anyone with an SBP greater than 140 received a score of 2. During the in-person exam, field staff measured height in cm from shoeless participants standing on uncarpeted floors and recorded weight to the nearest 0.1 kg. BMI was computed as kg/m2, and obesity is indicated by a BMI above 30. Those categorized as obese receive a score of 2, while those who are not receive a score of 0. Total cholesterol is categorized into deciles within each wave. Those with a total cholesterol measurement in the highest decile were given a score of 2, while those in the bottom 9 deciles were given a 0. Lastly, the physical activity score is derived from a series of survey questions about the frequency of participation in certain activities in the past week. Responses could range from 0 to 7 or more times (0-7). We added the number of times the respondent reported over all these questions. They were considered active if they participated in 2 or more active activities in the past week. The choice of 2x per week as a threshold was based on the attempt to harmonize with the original creation of the CAIDE score. Finally, APOE status was defined by having at least one ε4 allele (i.e., those with APOE ε2/ε4, ε3/ε4, or ε4/ε4 phenotypes).

APOE ε4: Saliva DNA collection occurred during Wave IV (96% consent rate). Archived samples (N=12,200) were eligible for genome-wide genotyping, with data available for N=9,974 participants after quality control procedures. Genomic data was further supplemented using DNA from venous blood collected during the Wave V biovisit for N=11,550 participants.

Blood Based Biomarkers: Values were log-transformed due to the skewness of biomarker concentration distributions. The values were then standardized to compare estimates on the same scale and account for different modes of sample collection between waves. A value of the limit of detection (LOD)/√2 was assigned to left-censored observations below the LOD. Of note, there was a high proportion of left-censoring for IL-10 in Wave IV (11.7%) and IL-1β in Wave V (29.8%). The development and validity of the assays used to assess inflammatory cytokines in DBS have been described elsewhere^1^. In some cases, instrument software provides extrapolated values below the LLOD if it can be differentiated from 0. Table S1 below shows the proportion of data that was below the LLOD’s and the proportion of data that was missing due to being below the LLOD (i.e. left-censored).

##### Table S1. Proportion of Left-Censoring by Biomarker

|  | **Biomarker** | **LLOD** | **N, % below LOD and extrapolated** | | **N, % missing due to left-censoring** | |
| --- | --- | --- | --- | --- | --- | --- |
| Wave IV (dried blood spots) | IL-6 | 0.4 | 356 | 9.0% | 130, | 3.0% |
|  | IL-8 | 0.2 | 0 | 0% | 10, | 0.3% |
|  | IL-10 | 0.4 | 358 | 10.3% | 476, | 11.7% |
|  | TNF-α | 0.9 | 5 | 0.1% | 24, | 0.6% |
| Wave V (venous blood) | IL-6 | 0.05 | 0 | 0% | 0 | 0% |
|  | IL-8 | 0.06 | 0 | 0% | 0 | 0% |
|  | IL-10 | 0.04 | 1 | 0.01% | 0 | 0% |
|  | TNF-α | 0.04 | 0 | 0% | 0 | 0% |
|  | IL-1β | 0.01 | 28 | 4.5% | 162 | 29.8% |
|  | NfL | 0.416 | 0 | 0% | 0 | 0% |
|  | Total Tau | 0.38 | 6 | 1.1% | 0 | 0% |

Cognitive Function: Immediate word recall involved participants being read a list of 15 words and repeating back as many as possible within 90 seconds. Delayed word recall required participants to recall as many words as possible from the same list of words after a few minutes, with 60 seconds to respond. Backward digit span consisted of FE’s reading a list of numbers and participants were asked to recite the span of numbers backwards, with the span length gradually increasing until an error occurred or a maximum of 7 digits. Participants were given two chances for each span length. All cognitive function scores were standardized into a z-score for analysis, with a higher score indicating higher memory cognition.

Social origins score: A factor score based on Wave I parental reports of education, occupation, household income, and household receipt of public assistance and was standardized to a z-score for analysis.

Recent inflammatory condition: This indicator measures included respondents who self-reported gum disease, active infection, injury, acute illness, and/or surgery in the past 4 weeks, and/or fever in the past 2 weeks. In Wave V, those who reported taking the following medications in the past four weeks were also included: cox-2 inhibitors, corticotrophines, glucocorticords, anti-rheumatics, anti-psoriatics, immunosuppressive agents or monoclonal antibodies. There was less specific information on medication in Wave IV so only those self-reporting inflammatory conditions were included.

Race/Ethnicity: We included the social construct of race and ethnicity as a covariate because there are documented racial and ethnic differentials in health, including conditions and biomarkers related to inflammation^2,3^, immunity^4^, cardiovascular health^5,6^, and disparities in Alzheimer’s Disease^7,8^. Therefore, disadvantaged minoritized groups are more likely to experience exposure to adverse environments and health care access inequities that are linked to both our biomarker exposures and cognitive function outcomes^9,10^. A race/ethnicity variable was constructed from the Wave V survey. Participants self-selected one or more boxes from a list including: “American Indian or Alaska Native”, “Asian”, “Black, African American”, “Hispanic”, “Pacific Islander”, “White”, and “Some other race or origin”. Those who chose one response were assigned to the corresponding category. Those who selected more than one option also answered a question on which category they most identified with and were assigned to that category. Anyone who skipped the question was assigned to the race/ethnicity they were categorized as in Wave I. Because some of the cell sizes for race and ethnicity were smaller than 10 in Wave V, some cell sizes were collapsed with others for masking and analyses.

### Sample Flowcharts

#### Supplementary Figure S1. Wave IV Sample Selection

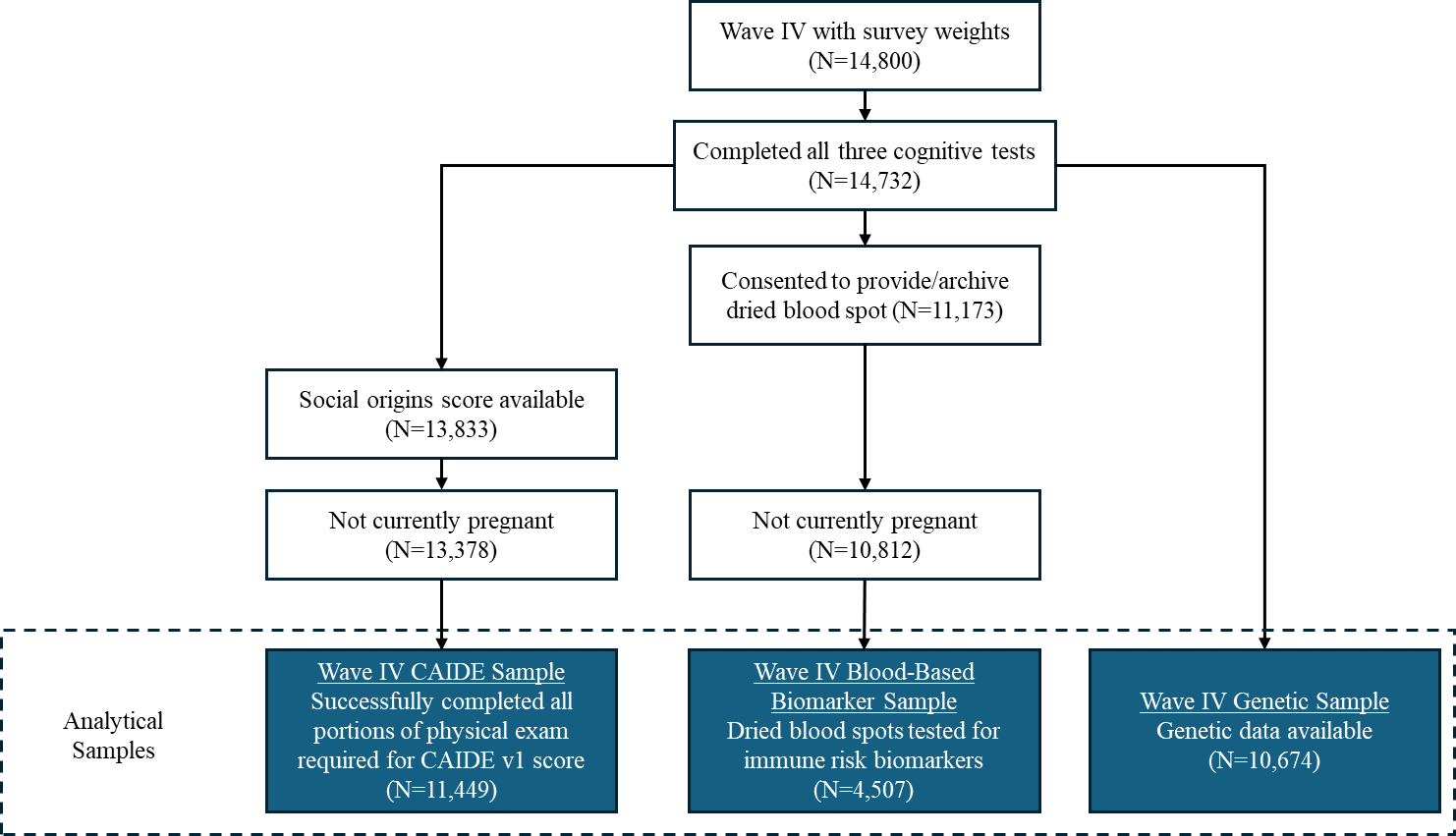

#### Supplementary Figure S2. Wave V Sample Selection

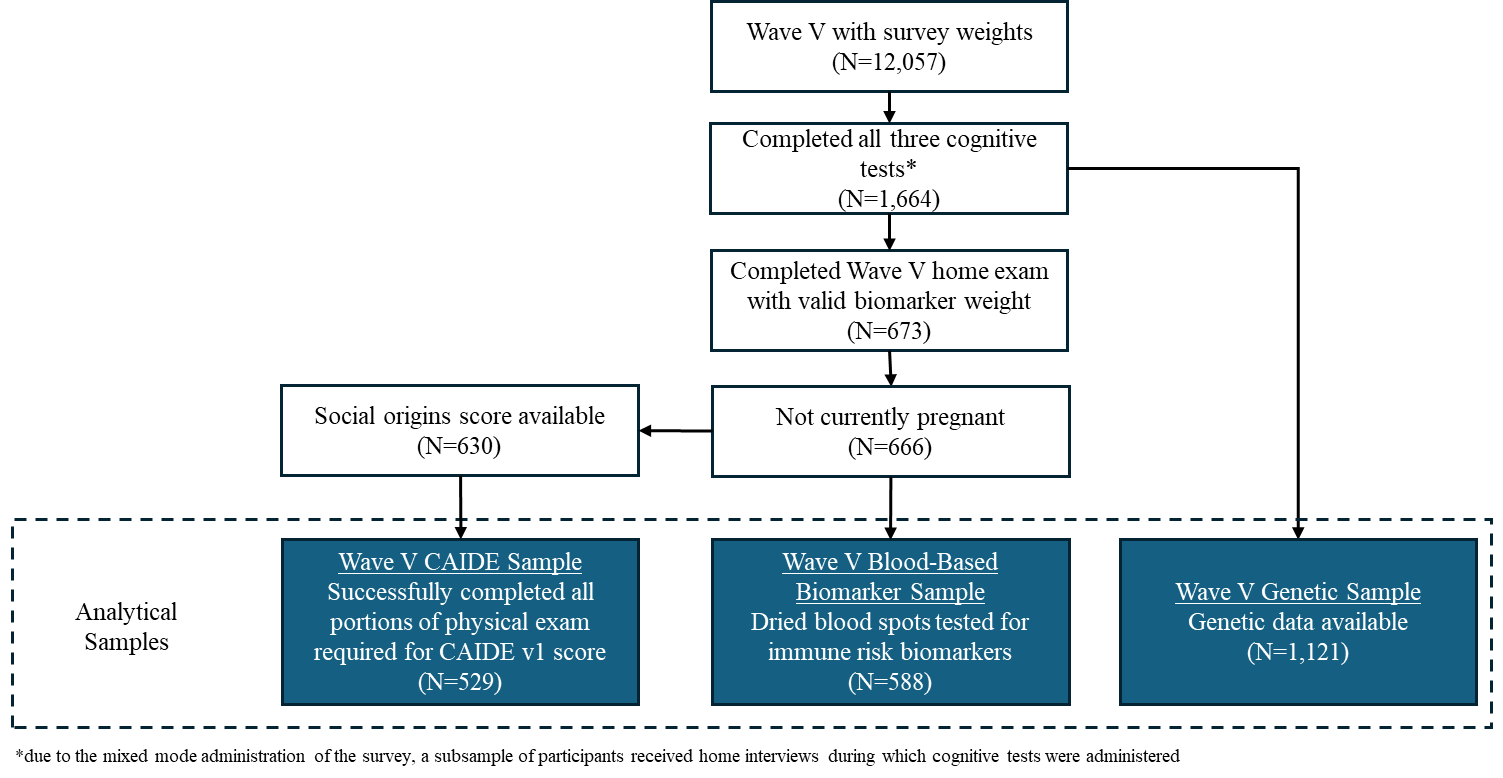

### Sample Characteristics of Wave IV overall and Wave V biosample

Given these data span two waves and different combinations of subsamples, the number of participants included in each analysis varies. We assume those with inflammatory cytokine data (N=5,019) in Wave IV consist of a random subsample of Wave IV overall. We also assume the major selection mechanism for inclusion in the Wave V analyses was participation in the Wave V biovisit. Descriptive statistics and analyses were weighted to account for sampling procedures, attrition and subpopulation analysis to make results generalizable to the U.S population of adults who were enrolled in middle school or high school in the 1994-1995 school year. To examine this assumption further, descriptive statistics of the overall Wave IV sample and the Wave V biosample is also included below in Table S2 for comparison to the analytic samples. In relation to Wave IV overall, each weighted sample was similar on demographics and other relevant variables. The only exception was a slightly lower proportion of participants with a college degree or higher in the immune risk factors sample (26% vs. 30%).  Compared to the Wave V biosample, the analytic samples had a lower proportion of females, a higher proportion of Black/African American participants, a lower proportion of those with a college degree or higher, a higher proportion of those with a recent inflammatory condition (17% vs. 14%), and higher median CAIDE scores. The samples were similar in other characteristics.

#### Table S2. Weighted Sample Characteristics compared to Overall Study Population, Add Health Waves IV-V

|  |  | **Wave IV Overall Sample** | | **Wave V Overall Sample** | |
| --- | --- | --- | --- | --- | --- |
|  |  | ***n=14,800*** | | ***n=5,269*** | |
| Age (years) | | 27.8 | (26.3, 29.3) | 37.4 | (35.9, 38.9) |
| Sex | |  |  |  |  |
|  | Female | 7870, | 49.32% | 3171, | 50.5% |
| Race | |  |  |  |  |
|  | American Indian or Alaska Native | 156, | 1.0% | 37, | 0.9% |
|  | Asian | 850, | 3.0% | 262, | 2.6% |
|  | Black | 3197, | 15.9% | 1035, | 17.3% |
|  | Hispanic | 2094, | 10.3% | 528, | 8.4% |
|  | Pacific Islander | 77, | 0.3% | 27, | 0.2% |
|  | Some other race or origin | 50, | 0.3% | 17, | 0.4% |
|  | White | 8376, | 69.2% | 3363, | 70.2% |
| Education | |  |  |  |  |
|  | College Degree or Higher | 4737, | 29.9% | 2449, | 41.3% |
|  | Some College and/or Technical Training | 6521, | 42.9% | 2023, | 40.7% |
|  | High School/GED or lower | 3538, | 27.2% | 796, | 18.1% |
| Recent Inflammatory Condition^a^ | | 2252, | 15.8% | 734, | 14.0% |
| Social Origins Score^b^ | | 0.1 | (-0.8, 1.0) | 0.3 | (-0.6, 1.1) |
| CAIDE score v1^c^ | | 5.3 | (3.2, 7.2) | 4.9 | (2.9, 6.9) |
| CAIDE score v2^c^ (with APOE status) | | 6.8 | (4.3, 9.1) | 6.2 | (3.8, 8.7) |
|  | *missing* | *5226* |  | *918* |  |
| Blood-based Biomarkers (log-transformed) | | *(n=5,019)* | |  |  |
|  | hsCRP (mg/L) | 0.7 | (-0.2, 1.7) | 0.6 | (-0.2, 1.5) |
|  | TNF-α (pg/mL) | 1.1 | (0.9, 1.3) | 0.9 | (0.7, 1.1) |
|  | IL-6 (pg/mL) | -0.2 | (-0.6, 0.3) | -0.4 | (-0.9, 0.1) |
|  | IL-10 (pg/mL) | -0.9 | (-1.4, -0.4) | -1.4 | (-1.8, -1.0) |
|  | IL-8 (pg/mL) | 4.3 | (4.0, 4.6) | 2.6 | (2.3, 3.1) |
|  | IL-1B (pg/mL) | NA |  | -3.7 | (5.0, -2.8) |
|  | NfL (pg/mL) | NA |  | 1.8 | (1.6, 2.1) |
|  | Total Tau (pg/mL) | NA |  | 0.8 | (0.4, 1.1) |
| APOE ε4 status | |  |  |  |  |
|  | ε4 carrier (ε2/ε4, ε3/ε4, or ε4/ε4) | 2957, | 27.4% | 1351, | 26.8% |
|  | *missing* | *4091* |  | *301* |  |
| Cognitive Function Tests | |  | | | |
|  | Immediate Word Recall | 6.0 | (4.7, 7.4) | 5.7 | (4.4, 7.0) |
|  | Delayed Word Recall | 4.6 | (3.3, 5.9) | 4.1 | (2.7, 5.5) |
|  | Backwards Digit Span | 3.4 | (2.4, 4.8) | 3.5 | (2.4, 4.9) |

Data are shown as N, survey-weighted % for categorical variables and weighted median, (IQR) for continuous variables.

^a^Indicator of any of the following conditions: gum disease, active infection, injury, acute illness, and/or surgery in the past 4 weeks, and/or fever in the past 2 weeks. Reported use of cox-2 inhibitors, corticotrophines, glucocorticords, anti-rheumatics, anti-psoriatics, immunosuppressive agents or monoclonal antibodies in the past 4 weeks included in Wave V

^b^Factor score based on Wave I parental reports of education, occupation, household income, and household receipt of public assistance (standardized to a z-score for analysis).

^c^Cardiovascular Risk Factors, Aging, and Incidence of Dementia (CAIDE) risk score version 1 is a weighted sum of education, sex, age, total cholesterol, systolic blood pressure, body mass index, and physical activity. Version 2 includes the same variables and adds APOE ε4 carrier status.

### Supplementary Analysis

To investigate how the strength of association between the CAIDE scores and cognitive function might vary at different life stages, we conducted the same cross-sectional linear regressions but restricted to the participants who were included in both waves (n=412 for CAIDE v1, n=406 for CAIDE v2 with APOE status included). Similarly, to assess how the association between APOE status and cognitive function might vary at different life stages, we completed linear regressions to assess the association between APOE status and Wave IV cognitive function and Wave V cognitive function among those with data in both waves (n=1063). Results are shown below in Figure S1.

#### Supplemental Figure S3: Associations between Inflammatory Biomarkers/CAIDE score and Cognitive Test Scores, Restricted to Participants with CAIDE scores and cognitive tests in both Wave IV and Wave V

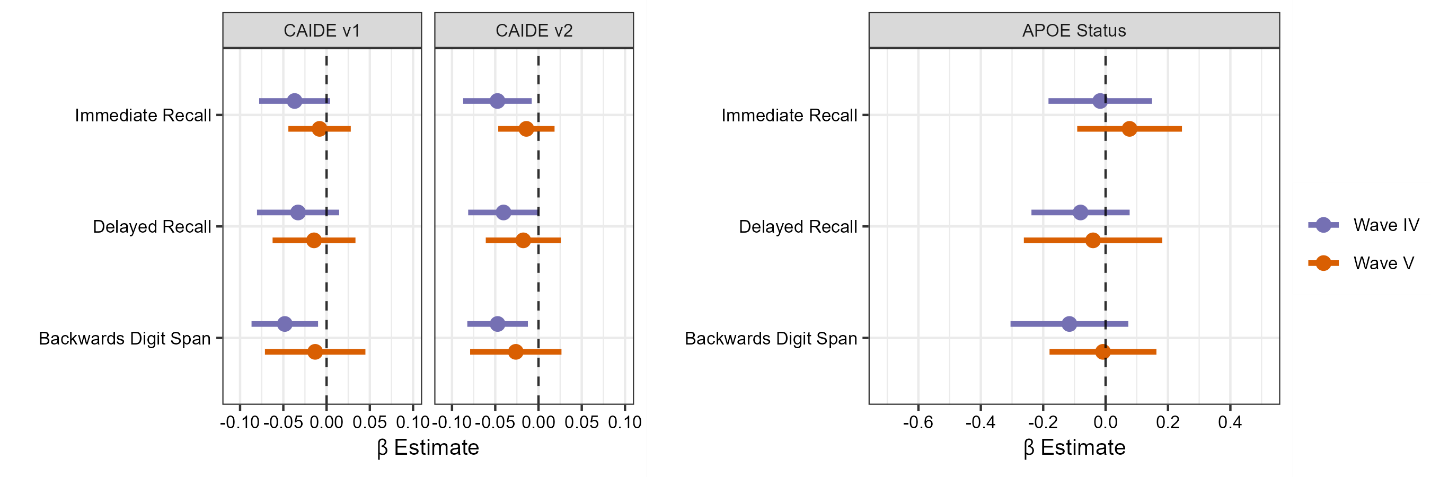

Each panel shows the β estimate and 95% CI for cross-sectional survey-weighted linear regressions where the CAIDE scores/APOE status (ε4 carrier vs. non-carrier) are the independent variable and cognitive function tests scores are the dependent variable.

CAIDE models adjusted for race/ethnicity, early life SES, and an indicator for inflammatory conditions. APOE models are adjusted for age and sex assigned at birth.

N=412 for CAIDE v1, N=406 for CAIDE v2, N=1063 for APOE status.

### Supplementary tables

β estimates and 95% confidence intervals for all analyses presented in Figures 1-4 can be found in the table below.

#### Supplementary Table S3. Associations Presented in Figures 1-4

| **Wave** | **Independent Variable** | **Cognitive Domain** | **N** | **Model** | **β Estimate** | **95% CI** | |
| --- | --- | --- | --- | --- | --- | --- | --- |
| IV | CAIDE v1 | Backwards Digit Span | 11449 | unadjusted | -0.05 | -0.06 | -0.04 |
| IV | CAIDE v1 | Backwards Digit Span | 11449 | adjusted | -0.03 | -0.04 | -0.02 |
| IV | CAIDE v1 | Delayed Recall | 11449 | unadjusted | -0.06 | -0.07 | -0.05 |
| IV | CAIDE v1 | Delayed Recall | 11449 | adjusted | -0.05 | -0.06 | -0.03 |
| IV | CAIDE v1 | Immediate Recall | 11449 | unadjusted | -0.06 | -0.07 | -0.05 |
| IV | CAIDE v1 | Immediate Recall | 11449 | adjusted | -0.05 | -0.06 | -0.04 |
| IV | CAIDEv2 | Backwards Digit Span | 8685 | unadjusted | -0.05 | -0.06 | -0.04 |
| IV | CAIDEv2 | Backwards Digit Span | 8685 | adjusted | -0.03 | -0.04 | -0.02 |
| IV | CAIDEv2 | Delayed Recall | 8685 | unadjusted | -0.05 | -0.06 | -0.04 |
| IV | CAIDEv2 | Delayed Recall | 8685 | adjusted | -0.04 | -0.05 | -0.03 |
| IV | CAIDEv2 | Immediate Recall | 8685 | unadjusted | -0.05 | -0.06 | -0.04 |
| IV | CAIDEv2 | Immediate Recall | 8685 | adjusted | -0.04 | -0.05 | -0.03 |
| V | CAIDE v1 | Backwards Digit Span | 529 | unadjusted | -0.04 | -0.09 | 0.01 |
| V | CAIDE v1 | Backwards Digit Span | 529 | adjusted | -0.03 | -0.08 | 0.02 |
| V | CAIDE v1 | Delayed Recall | 529 | unadjusted | -0.04 | -0.08 | 0.00 |
| V | CAIDE v1 | Delayed Recall | 529 | adjusted | -0.03 | -0.07 | 0.01 |
| V | CAIDE v1 | Immediate Recall | 529 | unadjusted | -0.03 | -0.06 | 0.01 |
| V | CAIDE v1 | Immediate Recall | 529 | adjusted | -0.02 | -0.06 | 0.02 |
| V | CAIDEv2 | Backwards Digit Span | 520 | unadjusted | -0.04 | -0.09 | 0.00 |
| V | CAIDEv2 | Backwards Digit Span | 520 | adjusted | -0.03 | -0.08 | 0.01 |
| V | CAIDEv2 | Delayed Recall | 520 | unadjusted | -0.03 | -0.07 | 0.00 |
| V | CAIDEv2 | Delayed Recall | 520 | adjusted | -0.02 | -0.06 | 0.02 |
| V | CAIDEv2 | Immediate Recall | 520 | unadjusted | -0.02 | -0.05 | 0.01 |
| V | CAIDEv2 | Immediate Recall | 520 | adjusted | -0.02 | -0.05 | 0.02 |
| IV | e4 carrier | Backwards Digit Span | 10674 | adjusted | -0.03 | -0.09 | 0.03 |
| IV | e4 carrier | Backwards Digit Span | 10674 | unadjusted | -0.03 | -0.09 | 0.03 |
| IV | e4 carrier | Delayed Recall | 10674 | adjusted | -0.02 | -0.07 | 0.04 |
| IV | e4 carrier | Delayed Recall | 10674 | unadjusted | -0.01 | -0.07 | 0.04 |
| IV | e4 carrier | Immediate Recall | 10674 | adjusted | -0.01 | -0.06 | 0.05 |
| IV | e4 carrier | Immediate Recall | 10674 | unadjusted | 0.00 | -0.06 | 0.05 |
| V | e4 carrier | Backwards Digit Span | 1121 | adjusted | 0.03 | -0.14 | 0.19 |
| V | e4 carrier | Backwards Digit Span | 1121 | unadjusted | 0.02 | -0.14 | 0.19 |
| V | e4 carrier | Delayed Recall | 1121 | adjusted | 0.00 | -0.22 | 0.21 |
| V | e4 carrier | Delayed Recall | 1121 | unadjusted | 0.00 | -0.21 | 0.22 |
| V | e4 carrier | Immediate Recall | 1121 | adjusted | 0.12 | -0.05 | 0.28 |
| V | e4 carrier | Immediate Recall | 1121 | adjusted | 0.12 | -0.04 | 0.29 |
| IV | e4e4 homozygote | Backwards Digit Span | 10674 | unadjusted | -0.06 | -0.25 | 0.12 |
| IV | e4e4 homozygote | Backwards Digit Span | 10674 | adjusted | -0.05 | -0.24 | 0.13 |
| IV | e4e4 homozygote | Delayed Recall | 10674 | unadjusted | -0.05 | -0.22 | 0.11 |
| IV | e4e4 homozygote | Delayed Recall | 10674 | adjusted | -0.02 | -0.19 | 0.14 |
| IV | e4e4 homozygote | Immediate Recall | 10674 | unadjusted | -0.09 | -0.26 | 0.09 |
| IV | e4e4 homozygote | Immediate Recall | 10674 | adjusted | -0.06 | -0.23 | 0.12 |
| V | e4e4 homozygote | Backwards Digit Span | 1121 | unadjusted | 0.09 | -0.48 | 0.66 |
| V | e4e4 homozygote | Backwards Digit Span | 1121 | adjusted | 0.08 | -0.48 | 0.65 |
| V | e4e4 homozygote | Delayed Recall | 1121 | unadjusted | -0.11 | -0.71 | 0.48 |
| V | e4e4 homozygote | Delayed Recall | 1121 | adjusted | -0.10 | -0.72 | 0.52 |
| V | e4e4 homozygote | Immediate Recall | 1121 | unadjusted | 0.21 | -0.16 | 0.58 |
| V | e4e4 homozygote | Immediate Recall | 1121 | adjusted | 0.22 | -0.15 | 0.59 |
| V | NfL | Backwards Digit Span | 588 | unadjusted | -0.08 | -0.17 | 0.00 |
| V | NfL | Backwards Digit Span | 588 | adjusted | -0.08 | -0.17 | 0.01 |
| V | NfL | Delayed Recall | 588 | unadjusted | 0.01 | -0.09 | 0.10 |
| V | NfL | Delayed Recall | 588 | adjusted | 0.01 | -0.10 | 0.12 |
| V | NfL | Immediate Recall | 588 | unadjusted | 0.01 | -0.09 | 0.11 |
| V | NfL | Immediate Recall | 588 | adjusted | 0.01 | -0.09 | 0.12 |
| V | total Tau | Backwards Digit Span | 584 | unadjusted | 0.06 | -0.06 | 0.18 |
| V | total Tau | Backwards Digit Span | 584 | adjusted | 0.04 | -0.08 | 0.16 |
| V | total Tau | Delayed Recall | 584 | unadjusted | -0.07 | -0.18 | 0.05 |
| V | total Tau | Delayed Recall | 584 | adjusted | -0.09 | -0.19 | 0.02 |
| V | total Tau | Immediate Recall | 584 | unadjusted | -0.13 | -0.24 | -0.02 |
| V | total Tau | Immediate Recall | 584 | adjusted | -0.14 | -0.24 | -0.04 |
| IV | IL-6 | Backwards Digit Span | 4507 | unadjusted | -0.08 | -0.13 | -0.04 |
| IV | IL-6 | Backwards Digit Span | 4507 | adjusted | -0.04 | -0.08 | 0.00 |
| IV | IL-6 | Delayed Recall | 4507 | unadjusted | -0.02 | -0.06 | 0.03 |
| IV | IL-6 | Delayed Recall | 4507 | adjusted | 0.00 | -0.04 | 0.05 |
| IV | IL-6 | Immediate Recall | 4507 | unadjusted | -0.02 | -0.07 | 0.02 |
| IV | IL-6 | Immediate Recall | 4507 | adjusted | -0.01 | -0.05 | 0.04 |
| IV | IL-10 | Backwards Digit Span | 4507 | unadjusted | 0.00 | -0.05 | 0.04 |
| IV | IL-10 | Backwards Digit Span | 4507 | adjusted | -0.01 | -0.06 | 0.03 |
| IV | IL-10 | Delayed Recall | 4507 | unadjusted | -0.03 | -0.08 | 0.01 |
| IV | IL-10 | Delayed Recall | 4507 | adjusted | -0.05 | -0.09 | -0.01 |
| IV | IL-10 | Immediate Recall | 4507 | unadjusted | 0.00 | -0.03 | 0.04 |
| IV | IL-10 | Immediate Recall | 4507 | adjusted | -0.01 | -0.04 | 0.02 |
| IV | IL-8 | Backwards Digit Span | 4507 | unadjusted | -0.05 | -0.11 | 0.00 |
| IV | IL-8 | Backwards Digit Span | 4507 | adjusted | -0.02 | -0.06 | 0.03 |
| IV | IL-8 | Delayed Recall | 4507 | unadjusted | -0.04 | -0.09 | 0.01 |
| IV | IL-8 | Delayed Recall | 4507 | adjusted | -0.01 | -0.06 | 0.03 |
| IV | IL-8 | Immediate Recall | 4507 | unadjusted | -0.04 | -0.09 | 0.00 |
| IV | IL-8 | Immediate Recall | 4507 | adjusted | -0.02 | -0.06 | 0.03 |
| IV | TNF-α | Backwards Digit Span | 4507 | unadjusted | -0.04 | -0.08 | 0.00 |
| IV | TNF-α | Backwards Digit Span | 4507 | adjusted | -0.03 | -0.07 | 0.01 |
| IV | TNF-α | Delayed Recall | 4507 | unadjusted | -0.02 | -0.06 | 0.02 |
| IV | TNF-α | Delayed Recall | 4507 | adjusted | -0.02 | -0.06 | 0.02 |
| IV | TNF-α | Immediate Recall | 4507 | unadjusted | -0.01 | -0.05 | 0.03 |
| IV | TNF-α | Immediate Recall | 4507 | adjusted | -0.01 | -0.04 | 0.03 |
| IV | hsCRP | Backwards Digit Span | 4485 | unadjusted | -0.08 | -0.12 | -0.04 |
| IV | hsCRP | Backwards Digit Span | 4485 | adjusted | -0.04 | -0.08 | 0.00 |
| IV | hsCRP | Delayed Recall | 4485 | unadjusted | 0.02 | -0.02 | 0.06 |
| IV | hsCRP | Delayed Recall | 4485 | adjusted | 0.03 | -0.01 | 0.06 |
| IV | hsCRP | Immediate Recall | 4485 | unadjusted | 0.00 | -0.04 | 0.03 |
| IV | hsCRP | Immediate Recall | 4485 | adjusted | 0.00 | -0.03 | 0.04 |
| V | hsCRP | Backwards Digit Span | 567 | unadjusted | -0.05 | -0.17 | 0.06 |
| V | hsCRP | Backwards Digit Span | 567 | adjusted | -0.01 | -0.13 | 0.11 |
| V | hsCRP | Delayed Recall | 567 | unadjusted | -0.02 | -0.12 | 0.09 |
| V | hsCRP | Delayed Recall | 567 | adjusted | 0.02 | -0.08 | 0.12 |
| V | hsCRP | Immediate Recall | 567 | unadjusted | -0.05 | -0.15 | 0.06 |
| V | hsCRP | Immediate Recall | 567 | adjusted | -0.02 | -0.13 | 0.09 |
| V | IL-6 | Backwards Digit Span | 588 | unadjusted | -0.15 | -0.23 | -0.06 |
| V | IL-6 | Backwards Digit Span | 588 | adjusted | -0.1 | -0.19 | -0.01 |
| V | IL-6 | Delayed Recall | 588 | unadjusted | -0.14 | -0.23 | -0.05 |
| V | IL-6 | Delayed Recall | 588 | adjusted | -0.09 | -0.18 | 0.00 |
| V | IL-6 | Immediate Recall | 588 | unadjusted | -0.05 | -0.17 | 0.06 |
| V | IL-6 | Immediate Recall | 588 | adjusted | -0.02 | -0.14 | 0.10 |
| V | IL-10 | Backwards Digit Span | 588 | unadjusted | -0.06 | -0.15 | 0.03 |
| V | IL-10 | Backwards Digit Span | 588 | adjusted | -0.05 | -0.14 | 0.04 |
| V | IL-10 | Delayed Recall | 588 | unadjusted | 0.00 | -0.12 | 0.12 |
| V | IL-10 | Delayed Recall | 588 | adjusted | 0.02 | -0.10 | 0.13 |
| V | IL-10 | Immediate Recall | 588 | unadjusted | -0.03 | -0.15 | 0.08 |
| V | IL-10 | Immediate Recall | 588 | adjusted | -0.02 | -0.13 | 0.10 |
| V | IL-8 | Backwards Digit Span | 588 | unadjusted | -0.19 | -0.30 | -0.09 |
| V | IL-8 | Backwards Digit Span | 588 | adjusted | -0.19 | -0.29 | -0.09 |
| V | IL-8 | Delayed Recall | 588 | unadjusted | -0.17 | -0.29 | -0.04 |
| V | IL-8 | Delayed Recall | 588 | adjusted | -0.17 | -0.29 | -0.05 |
| V | IL-8 | Immediate Recall | 588 | unadjusted | -0.18 | -0.27 | -0.09 |
| V | IL-8 | Immediate Recall | 588 | adjusted | -0.18 | -0.28 | -0.09 |
| V | TNF-α | Backwards Digit Span | 588 | unadjusted | -0.11 | -0.22 | -0.01 |
| V | TNF-α | Backwards Digit Span | 588 | adjusted | -0.12 | -0.25 | 0.01 |
| V | TNF-α | Delayed Recall | 588 | unadjusted | -0.05 | -0.17 | 0.07 |
| V | TNF-α | Delayed Recall | 588 | adjusted | -0.04 | -0.16 | 0.07 |
| V | TNF-α | Immediate Recall | 588 | unadjusted | -0.08 | -0.19 | 0.02 |
| V | TNF-α | Immediate Recall | 588 | adjusted | -0.07 | -0.19 | 0.04 |
| V | IL-1β | Backwards Digit Span | 588 | unadjusted | -0.11 | -0.24 | 0.01 |
| V | IL-1β | Backwards Digit Span | 588 | adjusted | -0.12 | -0.25 | 0.00 |
| V | IL-1β | Delayed Recall | 588 | unadjusted | -0.16 | -0.30 | -0.03 |
| V | IL-1β | Delayed Recall | 588 | adjusted | -0.15 | -0.28 | -0.03 |
| V | IL-1β | Immediate Recall | 588 | unadjusted | -0.12 | -0.22 | -0.01 |
| V | IL-1β | Immediate Recall | 588 | adjusted | -0.12 | -0.21 | -0.02 |

#### Supplementary Table S4. Associations Presented in Figure S1.

| **Wave** | **Independent Variable** | **Cognitive Domain** | **N** | **Model** | **Beta Estimate** | **95% CI** | |
| --- | --- | --- | --- | --- | --- | --- | --- |
| **CAIDE Scores** | |  |  |  |  |  |  |
| IV | CAIDE v1 | Backward Digit Span | 412 | unadjusted | -0.06 | -0.10 | -0.02 |
| IV | CAIDE v1 | Backward Digit Span | 412 | adjusted | -0.05 | -0.09 | -0.01 |
| V | CAIDE v1 | Backward Digit Span | 412 | unadjusted | -0.03 | -0.08 | 0.03 |
| V | CAIDE v1 | Backward Digit Span | 412 | adjusted | -0.01 | -0.07 | 0.04 |
| IV | CAIDE v1 | Delayed Recall | 412 | unadjusted | -0.05 | -0.10 | 0.00 |
| IV | CAIDE v1 | Delayed Recall | 412 | adjusted | -0.03 | -0.08 | 0.01 |
| V | CAIDE v1 | Delayed Recall | 412 | unadjusted | -0.02 | -0.07 | 0.02 |
| V | CAIDE v1 | Delayed Recall | 412 | adjusted | -0.01 | -0.06 | 0.03 |
| IV | CAIDE v1 | Immediate Recall | 412 | unadjusted | -0.05 | -0.09 | -0.01 |
| IV | CAIDE v1 | Immediate Recall | 412 | adjusted | -0.04 | -0.08 | 0.00 |
| V | CAIDE v1 | Immediate Recall | 412 | unadjusted | -0.01 | -0.05 | 0.03 |
| V | CAIDE v1 | Immediate Recall | 412 | adjusted | -0.01 | -0.04 | 0.03 |
| IV | CAIDE v2 | Backward Digit Span | 406 | unadjusted | -0.06 | -0.10 | -0.03 |
| IV | CAIDE v2 | Backward Digit Span | 406 | adjusted | -0.05 | -0.08 | -0.01 |
| V | CAIDE v2 | Backward Digit Span | 406 | unadjusted | -0.04 | -0.09 | 0.01 |
| V | CAIDE v2 | Backward Digit Span | 406 | adjusted | -0.03 | -0.08 | 0.03 |
| IV | CAIDE v2 | Delayed Recall | 406 | unadjusted | -0.06 | -0.10 | -0.02 |
| IV | CAIDE v2 | Delayed Recall | 406 | adjusted | -0.04 | -0.08 | 0.00 |
| V | CAIDE v2 | Delayed Recall | 406 | unadjusted | -0.03 | -0.07 | 0.01 |
| V | CAIDE v2 | Delayed Recall | 406 | adjusted | -0.02 | -0.06 | 0.03 |
| IV | CAIDE v2 | Immediate Recall | 406 | unadjusted | -0.06 | -0.11 | -0.02 |
| IV | CAIDE v2 | Immediate Recall | 406 | adjusted | -0.05 | -0.09 | -0.01 |
| V | CAIDE v2 | Immediate Recall | 406 | unadjusted | -0.02 | -0.05 | 0.02 |
| V | CAIDE v2 | Immediate Recall | 406 | adjusted | -0.01 | -0.05 | 0.02 |
| APOE ε4 | |  |  |  |  |  |  |
| IV | e4 carrier | Backward Digit Span | 1063 | unadjusted | -0.11 | -0.30 | 0.08 |
| IV | e4 carrier | Backward Digit Span | 1063 | adjusted | -0.12 | -0.30 | 0.07 |
| V | e4 carrier | Backward Digit Span | 1063 | unadjusted | 0.00 | -0.17 | 0.17 |
| V | e4 carrier | Backward Digit Span | 1063 | adjusted | -0.01 | -0.18 | 0.16 |
| IV | e4 carrier | Delayed Recall | 1063 | unadjusted | -0.08 | -0.25 | 0.08 |
| IV | e4 carrier | Delayed Recall | 1063 | adjusted | -0.08 | -0.24 | 0.08 |
| V | e4 carrier | Delayed Recall | 1063 | unadjusted | -0.05 | -0.27 | 0.18 |
| V | e4 carrier | Delayed Recall | 1063 | adjusted | -0.04 | -0.26 | 0.18 |
| IV | e4 carrier | Immediate Recall | 1063 | unadjusted | -0.02 | -0.19 | 0.15 |
| IV | e4 carrier | Immediate Recall | 1063 | adjusted | -0.02 | -0.18 | 0.15 |
| V | e4 carrier | Immediate Recall | 1063 | unadjusted | 0.07 | -0.10 | 0.24 |
| V | e4 carrier | Immediate Recall | 1063 | adjusted | 0.08 | -0.09 | 0.24 |
